# Healthy Japanese Dietary Pattern Is Associated with Slower Biological Aging in Older Men: WASEDA’S Health Study

**DOI:** 10.1101/2024.01.20.24300981

**Authors:** Takuji Kawamura, Mitsuru Higuchi, Tomoko Ito, Ryoko Kawakami, Chiyoko Usui, Kristen M. McGreevy, Steve Horvath, Radak Zsolt, Suguru Torii, Katsuhiko Suzuki, Kaori Ishii, Shizuo Sakamoto, Koichiro Oka, Isao Muraoka, Kumpei Tanisawa

## Abstract

Aging is the greatest risk factor for numerous diseases and mortality, and establishing geroprotective interventions targeting aging is required. Previous studies have suggested that healthy dietary patterns, such as the Mediterranean diet, are associated with delayed biological aging; however, these associations depend on nationality and sex. Therefore, this study aimed to investigate the relationship between dietary patterns identified through principal component analysis and biological aging in older men of Japan, one of the countries with the longest life expectancies. Principal component analysis identified two dietary patterns: a healthy Japanese dietary pattern and a Western-style dietary pattern. Eight epigenetic clocks, some of the most accurate aging biomarkers, were identified using DNA methylation data from whole-blood samples. Correlation analyses revealed that healthy Japanese dietary patterns were significantly negatively or positively correlated with multiple epigenetic age accelerations (AgeAccel), including AgeAccelGrim, FitAgeAccel, and age-adjusted DNAm-based telomere length (DNAmTLAdjAge). Conversely, the Western-style dietary pattern showed no significant correlation with any of the examined epigenetic AgeAccels or age-adjusted values. After adjusting for confounders, the healthy Japanese dietary pattern remained significantly negatively correlated with AgeAccelPheno and AgeAccelGrim and positively correlated with DNAmTLAdjAge. These findings suggest that a Western-style dietary pattern is not associated with biological aging, whereas a healthy Japanese dietary pattern is associated with delayed biological aging in older Japanese men. Our findings provide evidence that healthy dietary patterns may have beneficial effects on delayed biological aging in older Japanese men.

## 1 Introduction

Daily nutritional intake and diet are essential requirements for a healthy life, and overeating and malnutrition are closely associated with the development of metabolic, cardiovascular, and neurodegenerative diseases. Nutrient intake and diet are closely associated with aging and have been implicated in lowering the risk of age-related diseases and mortality (1). In recent years, an approach based on the “geroscience hypothesis” has been expanding, establishing intervention strategies that target the greatest risk factor for these diseases, aging itself, rather than addressing individual diseases (2, 3). From an economic perspective, it has been shown that a reduction in morbidity that improves health is more valuable than further increases in life expectancy and that targeting aging offers potentially larger economic gains than eradicating individual diseases (4). Given this background, research is underway to evaluate the efficacy of geroprotectants in delaying aging (5). However, evidence for the geroprotective effects of nutritional intake and diet is lacking.

Aging is a complex, multifaceted phenomenon influenced by genetic, environmental, and lifestyle factors and is characterized by a gradual decline in the physiological function and resilience of body systems over time. Epigenetic alterations have received increasing attention in recent years as hallmarks of aging, and epigenetic clocks based on age-related changes in DNA methylation patterns are promising biomarkers for testing the efficacy of geroprotective interventions (7). Most epigenetic clocks are calculated by (1) selecting key cytosine-phosphate-guanine (CpG) sites where hyper- and hypomethylation correlate with age and other phenotypes, weighting them with a linear model, and (2) creating an equation to estimate age based on the methylation level of each CpG site (8). Epigenetic clocks are associated with chronological age, onset of age-related diseases, and mortality (9,10) and are considered highly accurate biomarkers of an individual’s biological aging. Therefore, the epigenetic clock may be a useful biomarker for assessing the geroprotective effects of nutritional intake and diet.

Accumulating evidence suggests that the epigenetic clock is associated with lifestyle factors, such as body composition (11), physical activity (12, 13) and physical fitness levels (14, 15), smoking (16), and alcohol intake (16), and can be delayed or reversed by pharmacological and lifestyle interventions (17―20). It has also been suggested that healthy dietary patterns, diets rich in fruits and vegetables, and mediterranean diets may delay the epigenetic clocks (21―26). However, these studies have primarily focused on Western populations, and there is no literature available on Asians, who have a very different dietary culture, especially the Japanese, one of the longest-lived countries in the world. Considering that the Mediterranean diet rejuvenated the epigenetic clock in a nationality- and sex-specific manner in a limited nutritional intervention study (27), the relationship between dietary patterns and the epigenetic clock is expected to vary greatly depending on the characteristics of the study participants. We found that a rich intake of micronutrients, such as vitamins and minerals, was associated with delayed epigenetic clocks in older Japanese men, even after adjusting for body mass index (BMI) and smoking (15). However, when recommending geroprotective intervention strategies, it is necessary to investigate the relationship with the epigenetic clock, not on the basis of nutrient levels, but on a more comprehensive unit of dietary patterns.

Therefore, this study aimed to investigate the relationship between dietary patterns identified via principal component analysis (PCA) and epigenetic clocks. Specifically, we conducted a dietary survey in older men aged 65–72 years and identified two characteristic dietary patterns using PCA: a healthy Japanese dietary pattern and a Western-style dietary pattern. We also calculated Eight epigenetic clocks: Horvath clock, Hannum clock, BioAge4HAStatic, DNAmSkinBloodClock, DNAmPhenoAge, DNAmGrimAge, DNAmFitAge, and DNAm-based telomere length (DNAmTL) using the methylation data of DNA samples extracted from the whole blood of the participants. Through these surveys and measurements, we identified dietary patterns associated with delayed biological aging.

## 2. Methods

### 2.1 Participants

The study was conducted using the same procedures as in our previous study on participants of the Waseda Alumni’s Sports, Exercise, Daily Activity, Sedentariness, and Health Study (WASEDA Health Health Study) (15, 28—33). Briefly, this study included 169 men aged 65–72 years who participated in the baseline survey of Cohort D between March 2015 and March 2020 (see our previous study for details of the cohort (31)), 144 of whom were included in the order of measurement date from the earliest to the latest, after excluding those whose DNA sample quality did not meet the criteria (*n* = 11). The participants were briefed about the study and signed an informed consent form prior to the baseline survey. This study was approved by the Research Ethics Committee of Waseda University (approval numbers: 2014-G002 and 2018-G001) and conducted in accordance with the Declaration of Helsinki (1964).

### 2.2 Self-administered questionnaires and dietary assessment

Age (in years), smoking habits (current, former, and non-smoker), and frequency of alcohol consumption (less than once a week, 2–4 times a week, and ≥5 times a week) were investigated using a self-administered questionnaire. Dietary intake status was assessed using a brief self-administered dietary history questionnaire (BDHQ) using the same methods as in previous studies (28, 29, 32, 33). Briefly, the BDHQ consists of four pages and takes approximately 15 min to complete. After answering the questions, the examiner carefully checked to ensure there were no erroneous answers. Based on the Japanese Standard Tables of Food Composition (34), ad hoc computer algorithms for the BDHQ were used to estimate the dietary intakes of 58 food and beverage items, energy, and selected nutrients. The validity of the dietary intake data (energy, nutrients, and foods) assessed using the BDHQ was confirmed using 16-d semi-weighted dietary records as the gold standard (35, 36).

### 2.3 Anthropometric measurement

Height (cm) was measured using a stadiometer (YHS-200D; YAGAMI Inc., Nagoya, Japan). Body weight (kg) and fat content (%) were measured using a multifrequency bioelectrical impedance analyzer (MC-980A; Tanita, Tokyo, Japan) with light clothing and without shoes. BMI (kg/m^2^) was calculated using height and body weight measurements.

### 2.4 Blood sampling, DNA extraction, and measurement of epigenome-wide DNA methylation

The procedure from blood collection to sample storage has been described previously (15, 31). The participants were instructed to fast for at least 12 h the night before blood collection. Venous blood was collected from the forearm vein in a collection tube containing an anticoagulant (EDTA-2Na). DNA was extracted from whole blood using a QIAamp DNA Midi Kit (Qiagen, Germany) according to the manufacturer’s instructions. Extracted DNA was dissolved in Buffer AE (10 mM Tris-Cl, 0.5 mM EDTA, pH 9.0). Prior to DNA methylation measurements, DNA samples were adjusted to a concentration of ≥ 50 ng/μL and A260/280 purity in the range of 1.7–2.1. Epigenome-wide DNA methylation was measured using the same procedure as previously reported (13). Briefly, bisulfite conversion of genomic DNA was performed using the EZ DNA Methylation Kit (Zymo Research, Irvine, CA, USA), followed by hybridization using the Infinium MethylationEPIC BeadChip Kit (Illumina Inc., San Diego, CA, USA). Sample- and probe-based quality checks were performed using the minfi, meffil, and ewastools R packages (14). In this study, all 144 DNA samples met the criteria set by Illumina, and no samples were excluded.

### 2.5 Calculation of epigenetic clocks

Eight epigenetic clocks—Horvath clock, Hannum clock, BioAge4HAStatic, DNAmSkinBloodClock, DNAmPhenoAge, DNAmGrimAge, DNAmFitAge, and DNAmTL— and age acceleration or age-adjusted values for each of these were evaluated using the epigenome-wide DNA methylation data obtained as previously reported (22, 23, 37―42). The Horvath clock, also referred to as the pan-tissue clock, was developed as a multitissue age predictor (38), whereas the Hannum clock, BioAge4HAStatic, and DNAmSkinBloodClock were developed as blood-based age predictors (37, 39, 40). DNAmPhenoAge, DNAmGrimAge, DNAmFitAge, and DNAmTL were developed as predictors of morbidity and mortality and are also referred to as composite epigenetic clocks (22, 23, 41, 42). These four composite epigenetic clocks can predict biological status and age with higher accuracy than the four epigenetic clocks trained on chronological age alone (8).

### 2.6 Statistical analyses

To identify dietary patterns, we performed PCA based on energy-adjusted food intake using the density method for 52 food and beverage items, as previously described (28). More specifically, of the 58 items, six items (sugar added to coffee and black tea, three items usually added during cooking [salt, oil, and sugar], table salt and salt-containing seasoning at the table, and soup consumed with noodles) were excluded from the analysis (28). Two factors were retained based on the eigenvalue (>1), slope of the scree plot, and factor interpretability. As descriptive data, continuous variables are presented as mean ± standard deviation (SD) and categorical variables are presented as number of persons and percentages. The distributions of the two dietary pattern scores were confirmed using histogram plots, and as normality was not assumed, Spearman’s rank correlation analyses were performed to determine the correlation between each epigenetic age acceleration and the two identified dietary patterns. In parallel, because the two dietary pattern scores contained negative values, they were log-transformed after adding +5, followed by Pearson correlation and partial correlation analyses. A partial correlation analysis adjusted for BMI and smoking status was performed. Assuming a sample size of 144 and an *r* value of 0.235, the statistical power of Spearman’s correlation analysis was 0.800; assuming a sample size of 144 and an ρ value of 0.231, the statistical power of Pearson’s correlation analysis was 0.800. The statistical power of the partial correlation analysis was 0.800, assuming a sample size of 140 and *r* value of 0.236. All the statistical analyses were performed using SPSS version 26 (IBM Corporation, Chicago, IL, USA).

## 3 Results

### 3.1 Dietary patterns

In this study, two dietary patterns were identified using PCA (**Table 1**). The first factor was the “healthy Japanese dietary pattern,” which is characterized by a high intake of vegetables, fruits, soy products, and seafood. The second factor was named the “Western-style dietary pattern” based on the characteristics of a high intake of meat and processed meats, eggs, and mayonnaise dressings. These two dietary patterns accounted for 18.0% of the variance in food intake.

**Table 1.** Factor loading matrix for dietary patterns identified by the principal component analysis.

| **Food groups** | **Healthy Japanese dietary pattern** | **Western-style dietary pattern** |
| --- | --- | --- |
| Low fat Milk | 0.228 | -0.169 |
| Milk/yogurt | — | 0.173 |
| Chicken | — | — |
| Pork/beef | — | 0.441 |
| Hum/sausage/bacon | — | 0.593 |
| Liver | — | 0.153 |
| Squid/octopus/shrimps/shellfish | 0.443 | -0.169 |
| Small fish with bone | — | — |
| Canned tuna | — | 0.222 |
| Dried fish/salted fish | — | — |
| Oily fish | 0.377 | -0.383 |
| Lean fish | 0.152 | -0.182 |
| Egg | 0.218 | 0.374 |
| Tofu/atsuage† | 0.197 | — |
| Natto* | 0.590 | — |
| Potatoes | 0.178 | — |
| Pickled green leaves vegetable | 0.334 | 0.427 |
| Pickled other vegetables | 0.176 | 0.237 |
| Lettuces/cabbage (raw) | 0.609 | 0.353 |
| Green leaves vegetable | 0.744 | 0.265 |
| Cabbage/Chinese cabbage | 0.661 | 0.122 |
| Carrots/pumpkin | 0.838 | — |
| Tofu/atsuage† | 0.724 | -0.233 |
| Other root vegetables | 0.647 | — |
| Tomatoes | 0.295 | 0.530 |
| Mushrooms | 0.274 | -0.239 |
| Seaweeds | 0.574 | -0.478 |
| Western-type confectioneries | — | — |
| Japanese confectioneries | — | — |
| Rice crackers/rice cake/okonomiyaki‡ | — | -0.182 |
| Ice cream | — | 0.183 |
| Citrus fruit | — | -0.297 |
| Persimmons/strawberries/kiwifruit | 0.596 | -0.389 |
| Other fruit | 0.479 | -0.441 |
| Mayonnaise/dressing | — | 0.568 |
| Bread | -0.207 | — |
| Buckwheat noodles | -0.172 | -0.170 |
| Japanese noodles | -0.210 | -0.240 |
| Chinese noodle | -0.302 | — |
| Pasta | -0.271 | — |
| Green tea | 0.199 | — |
| Black tea/oolong tea | — | 0.369 |
| Coffee | — | — |
| Cola drink/soft drink | — | 0.189 |
| 100% fruit and vegetable juice | — | 0.227 |
| Rice | -0.251 | -0.331 |
| Miso soup | — | -0.194 |
| Sake | — | — |
| Beer | -0.323 | — |
| Shochu | -0.243 | — |
| Whisky | — | — |
| Wine | — | 0.162 |
| Variance explained | 11.155 | 6.857 |

A factor loading less than $\pm 0.15$ is represented by a dash for simplicity. \*: Fermented soybeans. †: Deep-fried tofu. ‡: Savoury pancake with various ingredients (meat, fish, and vegetable).

### 3.2 Characteristics of the participants

The predicted age values for each epigenetic clock were 62.2 ± 4.5 years for the Horvath clock, 55.1 ± 3.8 years for the Hannum clock, 52.3 ± 4.7 years for the BioAge4HAStatic, 65.7 ± 3.0 years for the DNAmSkinBloodClock, 56.7 ± 5.5 years for the DNAmPhenoAge, 69.2 ± 3.4 years for the DNAmGrimAge, 71.7 ± 3.5 years DNAmFitAge, and 6.6 ± 0.2 for DNAmTL, respectively. Age acceleration and age-adjusted values for each epigenetic clock are listed in **Table 2**.

**Table 2.**
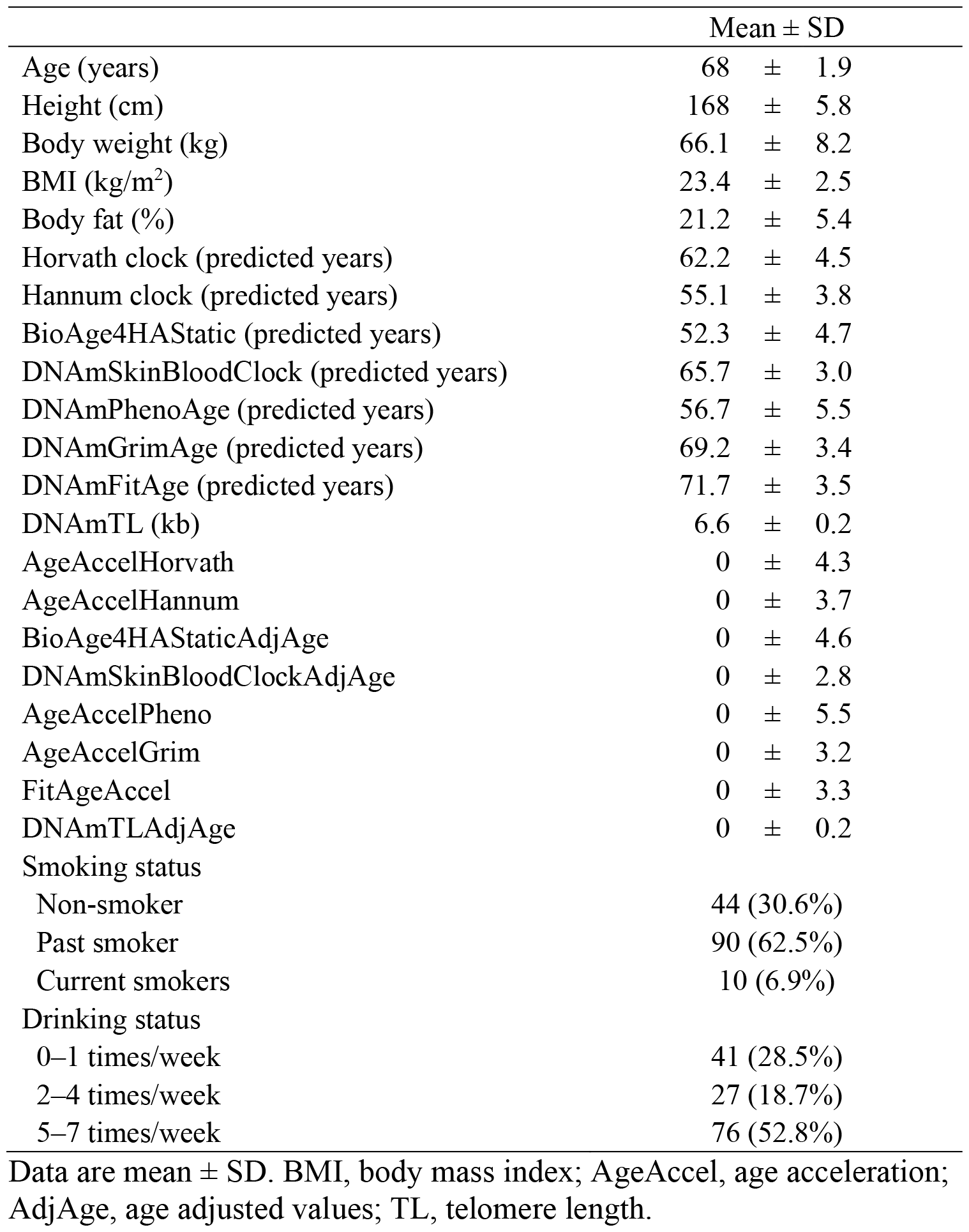
Characteristics of the participants (*n* =144).

### 3.3 Correlations between dietary pattern score and epigenetic clocks

**Figure 1**. shows the results of correlation analysis between the two dietary pattern scores and each epigenetic clock. For the healthy Japanese dietary pattern, both the actual and log-transformed values were significantly correlated with the composite epigenetic clocks. More specifically, healthy Japanese dietary pattern scores had significant negative correlations with AgeAccelGrim (ρ = -0.234; *p* = 0.005) and FitAgeAccel (ρ = -0.184; *p* = 0.027) and significant positive correlations with DNAmTLAdjAge (ρ = 0.197; *p* = 0.018) (correlation scatterplots are shown in **Figure S1**). The log-transformed healthy Japanese dietary pattern scores also had significant negative correlations with AgeAccelPheno (*r* = -0.173; *p* = 0.038), AgeAccelGrim (*r* = -0.266; *p* = 0.013), and FitAgeAccel (*r* = -0.181; *p* = 0.030) and significant positive correlations with DNAmTLAdjAge (*r* = 0.200; *p* = 0.016) (correlation scatterplots are shown in **Figure S2**). There were significant negative correlations between the log-transformed healthy Japanese dietary pattern scores and AgeAccelPheno (*r* = -0.177; *p* = 0.035) and AgeAccelGrim (*r* = -0.223; *p* = 0.008) and significant positive correlations with DNAmTLAdjAge (*r* = 0.205; *p* = 0.014), even after adjusting for BMI and smoking. In contrast, there were no significant correlations between healthy Japanese dietary patterns and the AgeAccelHorvath, AgeAccelHannum, BioAge4HAStaticAdjAge, or DNAmSkinBloodClockAdjAge scores. For the Western-style dietary pattern, neither the actual nor the log-transformed values were significantly correlated with any of the epigenetic clocks in any correlation analyses.

**Figure 1.**
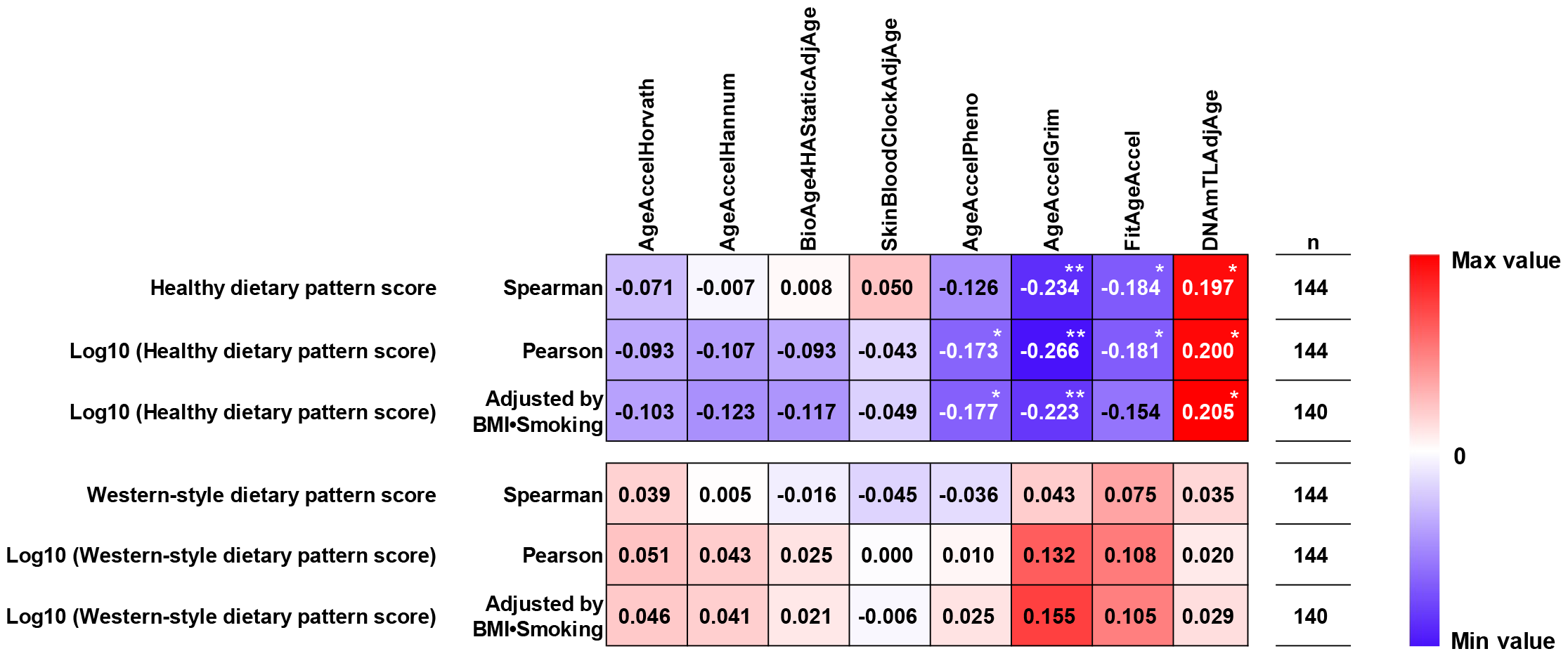
Correlations between dietary pattern score and epigenetic clocks. The values in the figure represent correlation and partial correlation coefficients. Partial correlation analysis was performed adjusted by body mass index (BMI) and smoking. Abbreviations: AgeAccel, age acceleration; AdjAge, age adjusted values; TL, telomere length. Significant correlations at *p* < 0.05 and *p* < 0.01 are indicated by * and **.

## 4 Discussion

This study revealed that a healthy Japanese dietary pattern exhibited significant negative or positive correlations with AgeAccelGrim, FitAgeAccel, and DNAmTLAdjAge scores in older Japanese men. Conversely, the Western-style dietary pattern showed no significant correlation with any of the examined epigenetic clocks. After adjusting for confounders, the healthy Japanese dietary pattern remained significantly negatively correlated with AgeAccelPheno and AgeAccelGrim and positively correlated with DNAmTLAdjAge. These findings suggest that a Western-style dietary pattern is not associated with biological aging, whereas a healthy Japanese dietary pattern is associated with delayed biological aging in older Japanese men.

Several previous studies have examined the relationship between dietary habits and the epigenetic clock based on individual dietary components, such as food items and nutrients. Collectively, the findings of these studies suggest a negative correlation between the intake of poultry, fish, vegetables, and fruit products and age acceleration and a positive correlation between red meat intake and age acceleration (21―23). In addition, studies have consistently shown that antioxidants in the blood are negatively correlated with accelerated age (21―23). Prior to the present study, we also found a negative correlation between estimated daily intake of vitamin C, *β*-carotene, iron, and copper and epigenetic age acceleration (15). However, previous research approaches lack a comprehensive understanding of the beneficial effects of daily dietary habits and lack specificity in establishing dietary guidelines and recommending dietary habit improvements.

To address this issue, Kresovich et al. (2022) (24) calculated scores for four dietary patterns (Dietary Approaches to Stop Hypertension diet, Healthy Eating Index-2015, Alternative Healthy Eating Index (aHEI-2010), and Alternative Mediterranean diet) based on the Block Food Frequency Questionnaire (Block FFQ) in 2694 women with an average age of 56 years and found that the scores for all four dietary patterns were negatively correlated with epigenetic age acceleration, most notably with PhenoAgeAccel and GrimAgeAccel. Of the four dietary patterns in this study, aHEL2010 had the strongest association with epigenetic age acceleration; similar results were reported in subsequent studies (16). Kim et al. (2022b) (25) also evaluated the Dietary Approaches to Stop Hypertension (DASH) score in 1995 using men and women with an average age of 67 years and found that higher scores were correlated with lower epigenetic age accelerations (*i*.*e*., Dunedin PoAm (43), PhenoAgeAccel, and GrimAgeAccel). Thomas et al. (2023) (26) reported a negative correlation between adherence to a Mediterranean diet and PhenoAgeAccel. Taken together, dietary patterns, such as aHEl-2010, DASH, and the Mediterranean diet, which are associated with a variety of diseases and mortality, appear to be associated with composite epigenetic clocks that can capture biological age.

Unlike previous studies based on dietary patterns, the present study identified two dietary patterns using PCA based on the results of the dietary assessment. These were the healthy Japanese dietary pattern and the Western-style dietary pattern, of which the healthy Japanese dietary pattern score was associated with epigenetic age acceleration of the composite epigenetic clocks that developed as predictors of morbidity and mortality in older Japanese men. The healthy Japanese dietary pattern is characterized by vegetables, fruits, seaweed, and natto (fermented soybeans) and is aligned to some extent with the dietary patterns used in previous studies, such as the aHEl-2010, DASH, and Mediterranean diets. Our findings indicate that a healthy Japanese dietary pattern delays biological aging processes and that the Western dietary pattern has no adverse effects on this process in older Japanese men. However, our cross-sectional study only found associations between dietary patterns and biological aging and could not determine a causal relationship between them.

Limited evidence from intervention studies suggests that combined lifestyle interventions, including dietary and exercise habits, reduce epigenetic age acceleration as compared to the control group (18, 19). Several other interventional studies have demonstrated that the effects of delayed epigenetic clock progression due to supplementation and altered dietary habits differ according to genotype, nationality, and sex (27, 44). The findings of the present study, based on the two categories of healthy Japanese- and Western-style dietary patterns, may provide important insights into establishing ideal dietary patterns for older Japanese men. Therefore, future longitudinal studies should follow both healthy Japanese dietary pattern scores and epigenetic age acceleration to clarify the causal relationship between these two factors in men and women. In addition, whether the healthy Japanese dietary patterns identified in this study would provide consistent results extending to populations of other nationalities and races should be tested.

## 5 Conclusion

We found that the Western-style dietary pattern was not associated with epigenetic age acceleration, whereas the healthy Japanese dietary pattern was associated with multiple composite epigenetic age accelerations that developed as predictors of morbidity and mortality in older Japanese men, suggesting that a healthy Japanese dietary pattern may have beneficial effects on delayed biological aging in older Japanese men.

## 7 Author Contributions

T.K., M.H., and K.T. conceptualized and designed the study. S.S., K.O., M.H., I. M., and K.T. conceived and supervised the study. T. K., T. I., R. K., C. U., K. S., K. I., S. S. and K. T. conducted the experiments. KM.M. and S.H. calculated epigenetic and epigenetic age accelerations. T.K. performed statistical analyses and created the figures. T. K., M. H., and K. T. interpreted the data. T.K. drafted the manuscript and all authors have read and agreed to its final version.

## 9 Acknowledgments

I would like to thank the staff of WASEDA’S Health Study and all the participants in the study. We also thank Editage for English editing (https://www.editage.jp/).

## 12 Data Availability Statement

The measurement data used to support the findings of this study are available from the corresponding author upon reasonable request.

## Supplementary data

**Figure S1.**
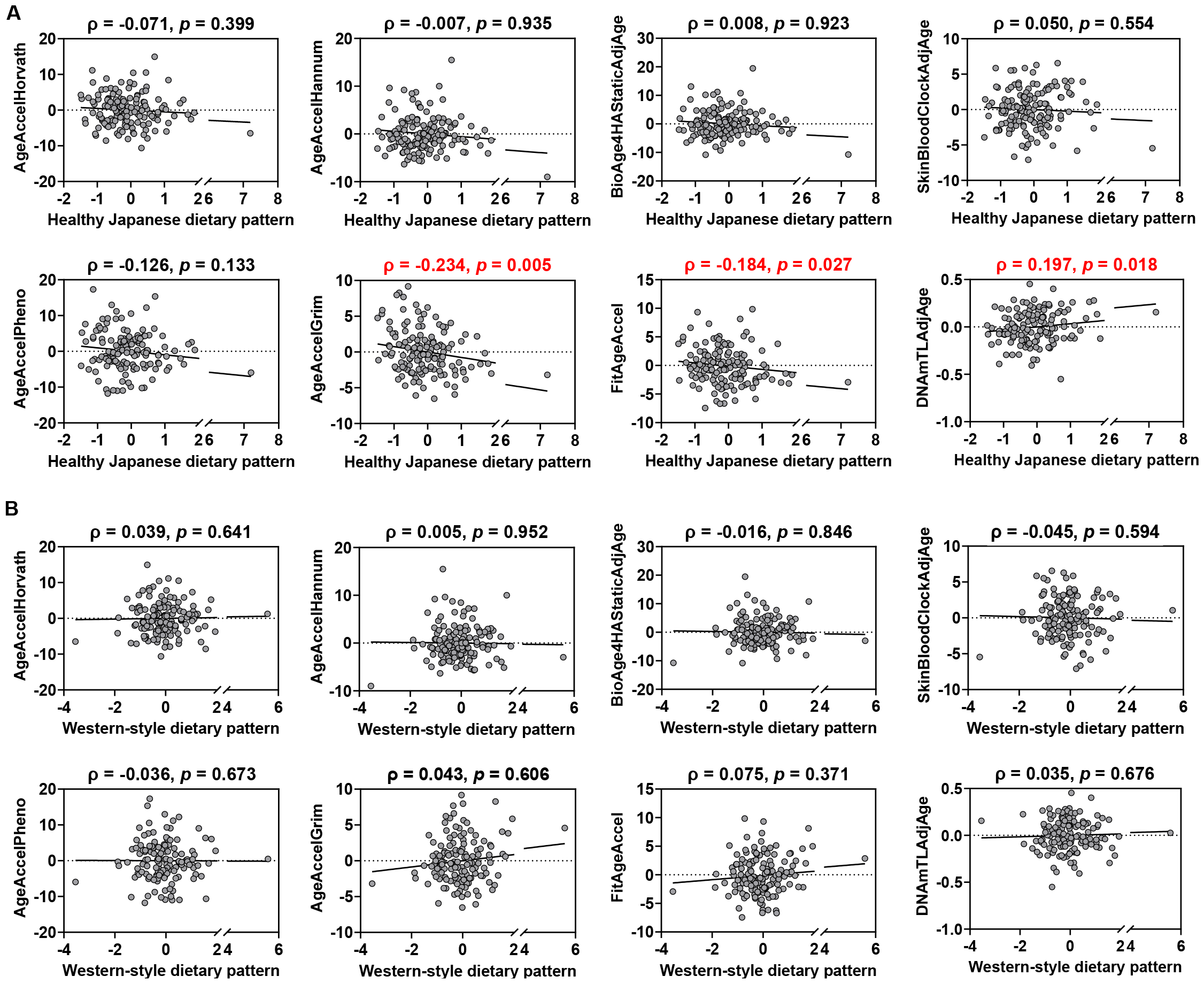
Spearman correlation scatterplots. **A)** Correlation between healthy Japanese dietary pattern scores and age acceleration (AgeAccel) or age adjusted (AdjAge) values for each epigenetic clock. **B)** Correlation between Western-style dietary pattern scores and AgeAccel or AdjAge. values for each epigenetic clock. Red letters indicate that the correlation is significant.

**Figure S2.**
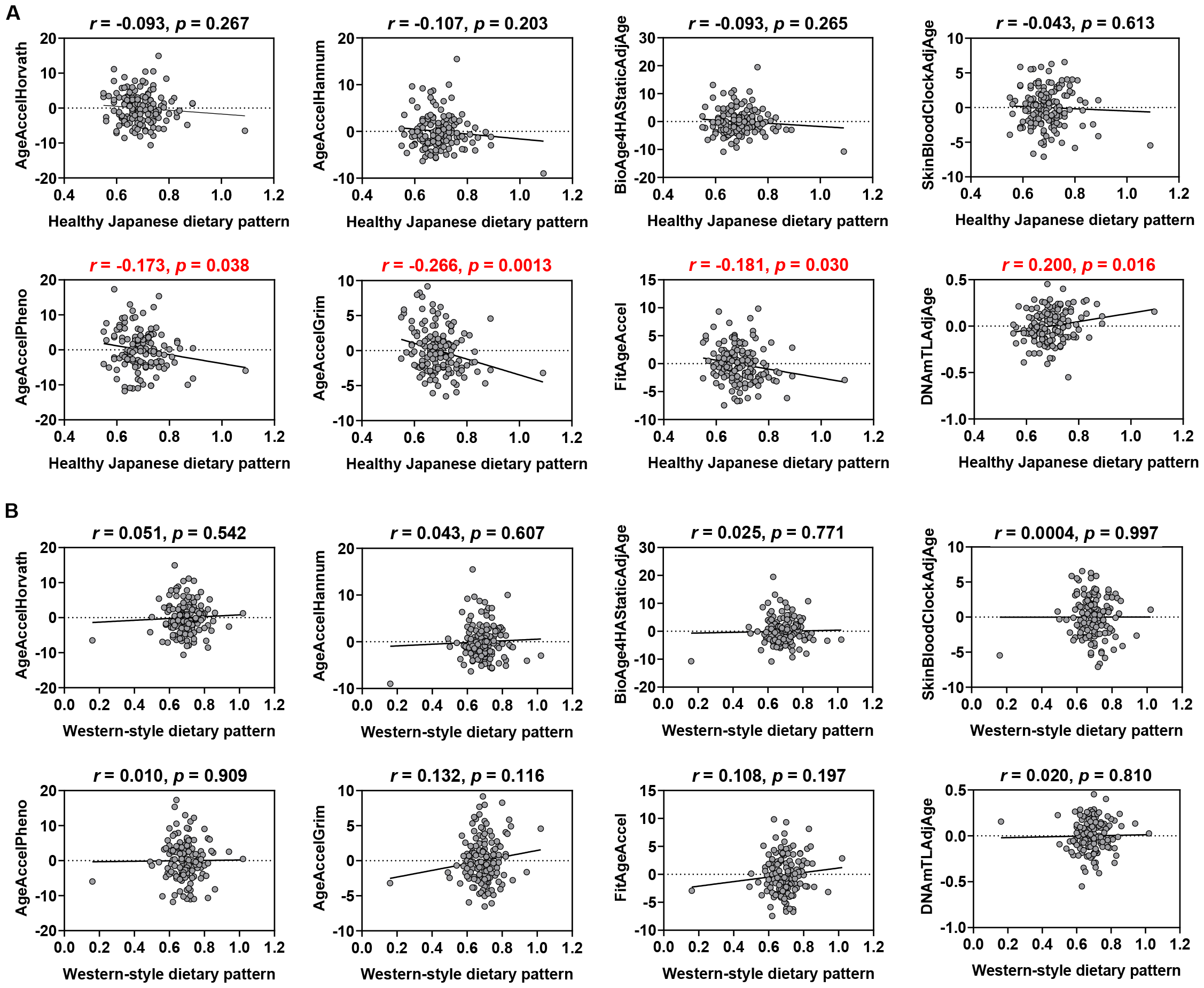
Pearson correlation scatterplots. **A)** Correlation between log10 (healthy Japanese dietary pattern scores) and age acceleration (AgeAccel) or age adjusted (AdjAge) values for each epigenetic clock. **B)** Correlation between log10 (western-style dietary pattern scores) and AgeAccel or AdjAge. values for each epigenetic clock. Red letters indicate that the correlation is significant.

